# National trends in suicides and male twin live births in the US, 2003 to 2019: an updated test of collective optimism and selection *in utero*

**DOI:** 10.1101/2023.07.21.23293010

**Authors:** Parvati Singh, Samantha Gailey, Abhery Das, Tim A. Bruckner

**Author notes:** **Author contributions:** PS conceived the study, acquired data, conducted data analyses and developed the manuscript; SG assisted with data acquisition and manuscript preparation; AD contributed to manuscript preparation and review; TAB conducted analyses and supervised manuscript development and review.

## Abstract

Prior research based on Swedish data suggests that collective optimism, as measured by monthly incidence of suicides, correlates inversely with selection *in utero* against male twins in a population. We test this finding in the US, which reports the highest suicide rate of all high-income countries, and examine whether monthly changes in overall suicides precede changes in the ratio of male twin to male singleton live births. Consistent with prior work, we also examine as a key independent variable, suicides among women aged 15-49 years. We retrieved monthly data on suicides and the ratio of male twin to singleton live births from CDC WONDER, 2003 to 2019, and applied Box-Jenkins iterative time-series routines to detect and remove autocorrelation from both series. Results indicate that a one percent increase in monthly change in overall suicides precedes a 0.005 unit decline in male twin live births ratio 6 months later (coefficient = -0.005, *P* value = 0.004). Results remain robust to use of suicides among reproductive-aged women as the independent variable (coefficient = -0.0012, *P* value = 0.014). Our study lends external validity to prior research and supports the notion that a decline in collective optimism corresponds with greater selection *in utero*.

## Introduction

The US reports the highest suicide rate (14 deaths per 100,000 people) among all high-income countries. Experts attribute this trend to increased national despair, that may approximate the bleak social and economic outlook experienced by a society on average^1^. Not surprisingly, suicides appear to track or correlate positively with some aggregate indicators of national despair ^2–7^.

In his seminal treatise on social determinants of suicides, Durkheim (1897) posited that suicide prevalence in a population is a consequence of the underlying social structure, beliefs and practices that he referred to as *collective inclination* ^8^. A population’s *collective inclination* towards suicide may exert coercive pressure on individuals towards suicide completion and may vary based on social despair during stressful periods ^8^. As an extreme consequence of increased despair, trends in suicides may reflect perturbations in aggregate sentiments of despair across place and time ^6, 7, 9^. Ecologically, collective despair or (conversely) optimism may reflect a population’s shared outlook ^10^ and manifest as contagious, collective expectations about the future ^11–14^. Suicides, by definition, reflect termination of the future. Whereas research on suicides and optimism largely focuses on individual-level mechanisms, national trends in select measures of optimism appear to explain (to an extent) spatial and temporal variation in suicides ^2–7^. Collective optimism, however, remains a complex construct that may derive not just from aggregate economic expectations, but also social integration, cohesion and social capital ^15–19^. Quantification of collective optimism in a manner that accounts for all facets of this construct remains difficult. However, as suggested by Durkheim, to the extent that suicides correspond with a society’s collective experiences of despair and future outlook, the patterning of suicides in a population may signal periods of acute decline in collective optimism ^8^.

Decline in collective optimism may also manifest as risk aversion or reduced willingness to invest in high-cost, uncertain ventures ^20^. Uncertainty and unfavorable expectations about the future correspond with reduction or postponement of expensive, long-term investments such as fertility, home purchase or new business creation ^15, 20, 21^. Childbearing remains one of the most fundamental, long-term investments over the life course. Substantial research documents reproductive risk-aversion during stressful ambient circumstances^22^ that, in turn, may also correspond with reduced collective optimism. Parental and social optimism appears to vary inversely with risk aversion, which spills over into conscious and non-conscious decisions regarding offspring conception, sex-specific fetal loss and fitness of resulting live births ^23, 24^.

Selection *in utero*, defined as spontaneous abortion of gestations (also referred to as fetal loss), offers one mechanism through which populations may respond to heightened uncertainty, or reduced collective optimism. ^22^. Selection *in utero* against frail or risky gestations, by way of increased spontaneous abortion or fetal loss, may indicate a pregnant person’s non-conscious, biological risk-averse response to unfavorable external circumstances. These unfavorable circumstances may diminish a frail infant’s survival or reproductive success should the pregnancy result in a live birth ^24^. Relative to females, male gestations appear particularly vulnerable to selection *in utero* owing to their greater demand on maternal caregiving resources but lower likelihood of yielding grandchildren (i.e. reproductive success) if born in stressful conditions ^22, 24^. Male twin gestations, on average, occupy the right tail of the gestational frailty distribution and fare poorer in terms of relative survival, longevity and (future) reproductive success relative to all other types of gestations ^25, 26^. Hence, the incidence of male twin live births (or *selection in utero* against male twins) may indicate the extent to which a population is willing to invest in risky gestations that, in turn, may gauge a population’s sensitivity to variations in collective optimism ^13, 21, 27^

Collective optimism may serve as a shared precursor of both (a) increase in suicides and (2) decline in male twin live births. If a population exhibits risk-aversion following sudden perturbations in collective optimism, temporal changes in one of the most acute indicators of despair-suicides-should precede changes in population-level markers of *selection in utero*, such as the incidence of male twin live births ^1, 13^. Two recent studies support this hypothesis. Karasek et al. (2015) report an inverse association between a measure of consumer confidence index among reproductive aged women (indicative of macroeconomic climate) and male twin births in Sweden ^21^. This research suggests that women may ‘sense’ ambient macroeconomic disturbances and, as a risk-averse response, yield fewer-than-expected male twins in the short-term (two months following extreme decline in consumer confidence). Catalano et al. (2020) test this phenomena through the lens of collective optimism and examine the relation between suicides among 15-49 years old women and male twin births in Sweden ^13^. They find an inverse association between female suicides and male twinning in that greater-than-expected incidence of suicide among reproductive aged women precedes a decline in male twin live births by three months ^13^. To our knowledge, no other studies that have examined the relation between collective optimism and male twin live births.

Replication and extension of this work to other national contexts may hold interest among evolutionary theorists, fertility scholars, and epidemiologists. The external validity of findings from tests of collective optimism in Sweden to other populations remains unexplored. Sweden differs markedly from the US in terms of healthcare systems, social safety nets, racial/ethnic diversity and income inequality ^31, 32^. We examine whether and to what extent sudden variations in collective optimism, as indicated by monthly changes in overall suicides, precede changes in the ratio of male twin to male singleton live births in the US using publicly available, nationally aggregated monthly data from 2003 to 2019. Consistent with prior work, we also test this relation with suicides among women aged 15-49 years ^13, 21^.

## Methods

### Data

We retrieved data on monthly counts of (1) male twin and singleton live births and suicides (mortality from intentional self-harm, ICD-10 Codes: X60-X84) from the Centers for Disease Control and Prevention’s online national database-CDC WONDER-for the period of 2003 to 2019 ^33–35^. CDC WONDER reports data from the National Center for Health Statistics’ Division of Vital Statistics and provides national, regional and temporal aggregates by select subgroups (e.g. gender, age) and health conditions ^33–35^. These data are publicly available and are extensively utilized by epidemiologists worldwide ^36^. We selected the time period of January 2003 to December 2019 because monthly counts of twin births (by sex) for the US are not available in CDC WONDER prior to 2003. We excluded the year 2020 from our analysis to limit confounding by the COVID-19 pandemic. These restrictions yielded four nationally aggregated data series of 204 months each: (1) male twin live births, (2) male singleton live births, (3) overall suicides (all ages, all sexes) and (4) suicides among women aged 15-49 years. This study was deemed exempt from IRB review as we used publicly available, aggregate, de-identified data.

### Variables

We defined as our outcome the ratio of male twins to male singleton live births per month. It is plausible that a general decline in male births may directly reduce the number of male twin births as well. For this reason, we used the monthly counts of male twin and male singleton live births to develop a monthly male twin ratio (male twin live births/male singleton live births) series. Our outcome formulation differs from that of prior research that utilizes the odds ratio of male twins relative to female twin births ^13^. Research on parental optimism and adaptive mechanisms with respect to sex ratios at birth reports an inverse, lagged relation between male and female twin live births ^23, 37^. Put simply, a decline in live-born male twins may precede an increase in female twin live births (and vice versa) ^37^. We thus contend that the formulation of odds ratio of male twin births, as utilized by Catalano et al (2020), may distort the temporal lag and/or the magnitude of observed associations owing to endogenous, inverse relations between the numerator (odds of male twin births) and denominator (odds of female twin births) of the outcome.

As our independent variable we used overall suicides, across all age and sex groups, per month. Overall suicides gauge several common underlying ambient risk factors shared across age and sex groups such as economic inequality and access to mental health care. Consistent with prior work, we also examine the robustness of results to suicides among women aged 15-49 years. This population may show heightened sensitivity to the social environment ^13, 21^.

We modelled sudden changes in collective optimism (or ‘shocks’) by converting monthly counts of suicides into percent monthly change in suicides. We subtracted the observed count of suicides in the previous month (x_m-1_) from those in a given month (x_m_) and divided this difference by the previous month’s observation (i.e. [x_m_- x_m-1_]/x_m-1_). This transformation offers the dual advantage of (1) centering around zero the strong, upward trend in suicides in the US documented in prior research ^38^ and (2) yielding sudden variations (i.e. volatility rather than levels) depending on whether suicides increased (positive) or decreased (negative) relative to the previous month. Prior research examining population-level impact of ecological stressors on perinatal and psychiatric outcomes similarly uses change scores when gauging ambient ‘shocks’ ^39, 40^.

### Analysis

We aim to test whether greater-than-expected change in monthly incidence of overall suicides in the US precedes a reduction in male twins born in the population. We also test this relation with respect to suicides among women aged 15-49 years. Our outcome series, however, may exhibit temporal patterns such as seasonality, trends, and persistence of “memory” from a preceding month into following months. Because of this patterning, also known as autocorrelation, the expected value of the outcome variable (in a given month) may not equal the mean of past values ^41, 42^. This circumstance violates the assumptions of correlational tests ^41, 42^. Analysis of autocorrelated data, in such cases, may yield spurious relations between the exposure and outcome owing to non-independence of outcome observations (violation of the i.i.d. assumption) and non-zero mean of residuals ^41, 42^.

We controlled for autocorrelation by modeling the expected value of the outcome series as a function of its past values. We performed this modeling using Autoregressive, Integrated Moving Average (ARIMA) time-series approach wherein we applied iterative pattern recognition routines developed by Box & Jenkins (2015) to detect autocorrelation parameters and control for them in our analysis ^43^. Epidemiologists routinely use ARIMA time-series analytic methods in longitudinal research on the relation between ambient stressors and birth outcomes ^13, 21, 37^. Identification of autocorrelation parameters allows for prediction of counterfactuals—that is, the monthly series of male twin ratio based solely on its inherent patterns, in the absence of potential perturbations induced by changes in the exposure variable. This counterfactual series thus contains expected values of the outcome under the null hypothesis ^41–43^. Removal of autocorrelation in the outcome also yields normally distributed residuals with a mean of zero ^41–43^.

We conducted ARIMA analysis using software from Scientific Computing Associates (SCA) ^44^. Our analytic steps appear below:

1. We used Box-Jenkins time-series methods to identify and remove autocorrelation in the monthly series of male twin ratio ^43^.
2. We applied the Box-Jenkins routines to identify and remove autocorrelation in the percent monthly change in overall suicides series ^43^. This exercise yielded exposure residuals that align with the classic correlational test, dating back to Fisher, wherein the residuals indicate deviation from expected values of the series, net of autocorrelation ^45^. We defined this residualized series as our exposure or independent variable.
3. We applied the exposure residuals obtained in step 2 to the ARIMA model devised in step 1. We specified exposure lags of 2, 3, 4, 5, 6 months based on prior work, the average gestational age of live-born male twins in the US (∼34 to 37 weeks) ^34^, and the hypothesized gestational ages at which male twins *in utero* appear sensitive to ambient stress ^13, 21^.
4. We inspected the residuals obtained from the time-series model in step 3 for autocorrelation. If any were found, we inserted relevant ARIMA parameters into the error term.
5. We conducted a sensitivity test by repeating step 3 using log transformed (natural logarithm) male twin ratio series as the outcome to gauge whether results from step 3 were sensitive to heteroskedastic variance and influential outliers.
6. As a robustness check of whether our findings align with prior research, we repeated steps 2 and 3 with de-trended residuals of percent monthly change in suicides among females aged 15-49 years as the exposure.

## Results

Table 1 presents the descriptive statistics of our analytic data. The 204 months in our time-series analysis yielded a total of 1,129,712 male twins and 33,850,336 male singleton births. Total number of overall suicides over our study period equaled 670,893, with 73,826 suicides among females aged 15-49 years. There were approximately 5,538 male twin births per month (standard deviation = 327.08) and male twin ratio per month averaged 0.0334 (standard deviation = 0.001). Percent monthly change in overall suicides averaged 0.0045 (standard deviation = 0.071) and among females aged 15-49 years, averaged 0.0075 (standard deviation = 0.107).

**Table 1:** Counts and distribution of male twin live births, male singleton live births, male twin ratio, overall suicides and suicides among females aged 15 to 49 years in the US, from 2003 to 2019.

| Variables | Descriptive statistics |
| --- | --- |
| Total number of male twin live births | 1,129,712 |
| Total number of male singleton live births | 33,850,336 |
| Mean monthly count of male twin live births (Standard deviation) | 5537.81 (327.08) |
| Mean monthly male twins ratio (Standard deviation) | 0.0334 (0.001) |
| Total number of overall suicides | 670,893 |
| Mean percent monthly change in overall suicides (Standard deviation) | 0.0045 (0.071) |
| Total number of suicides among females aged 15-49 years | 73,826 |
| Mean percent monthly change in suicides among females aged 15-49 years (Standard deviation) | 0.0075 (0.107) |

Figure 1 plots the monthly male twin ratio from January 2003 to December 2019. We observe a slight increase in this ratio starting in 2009 that persists through 2019 (Figure 1).

**Figure 1:**
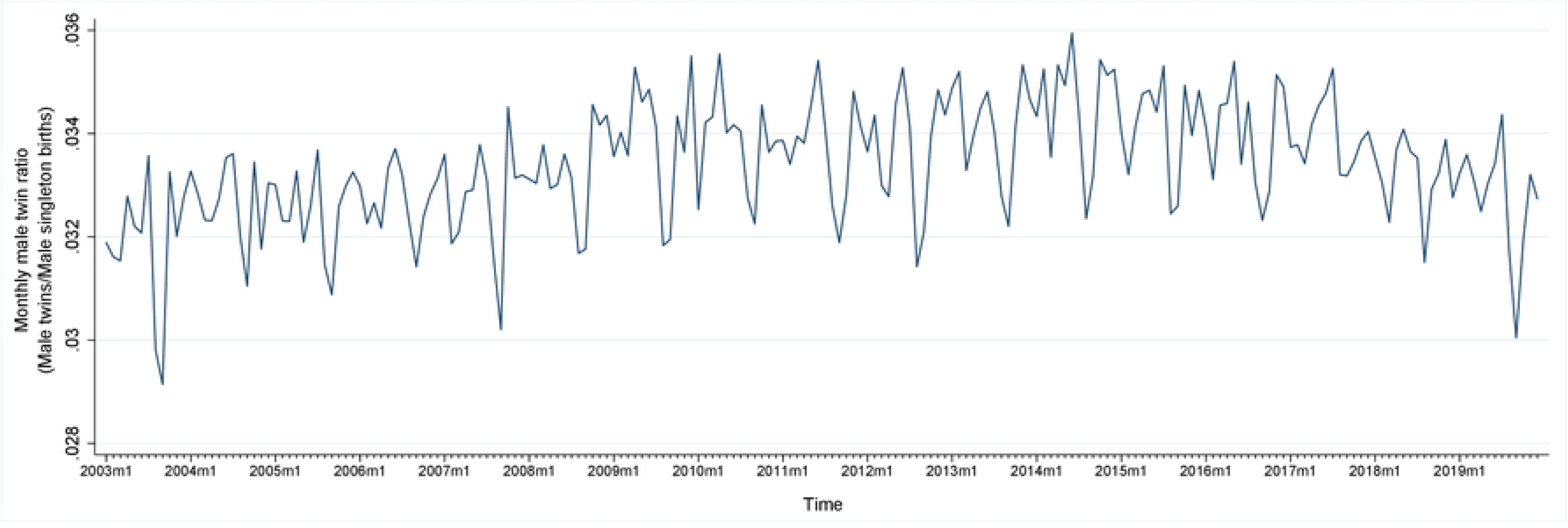
Plot of monthly male twin ratio (male twin live births/male singleton live births) in the US, from January 2003 to December 2019.

Figure 2 presents the percent monthly change in overall suicides. As expected from our transformation of monthly suicide counts into percent monthly change, this series centers around zero but shows seasonal peaks and troughs (Figure 2).

**Figure 2:**
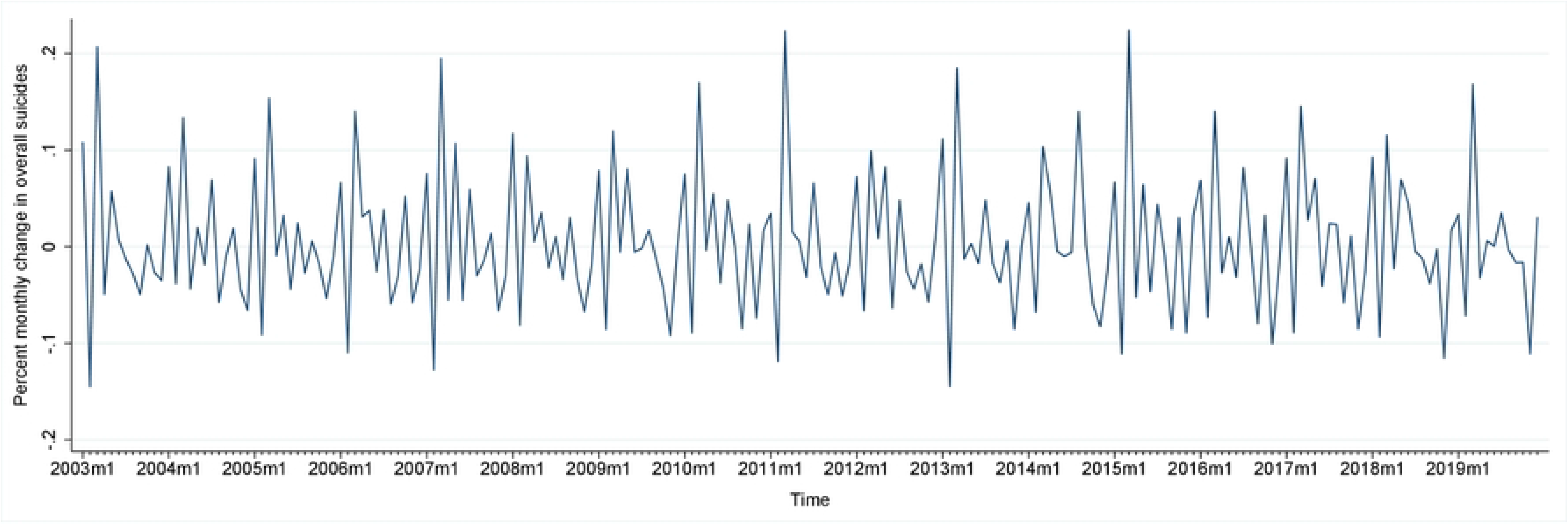
Plot of percent monthly change in overall suicides in the US, from January 2003 to December 2019.

The plot of percent monthly change in female suicides (aged 15-49 years) appears in Supplement Figure 1.

Box-Jenkins routines identified autoregression (i.e. long-term retention or ‘echoes’ of observations) in the monthly series of male twin ratios at lags 1 and 12 months (AR 1, 12). Figure 3a plots the residuals of monthly male twin ratios after removal of AR 1, 12 from the original series and represents the counterfactual. Application of Box-Jenkins ARIMA routines to percent monthly change in overall suicides detected two moving average parameters at lags 1 and 12 indicating high or low values of residual errors being remembered at 1 and 12 months later. We also detected an integration parameter that required differencing of the percent monthly change in overall suicides series over the previous 12 months (I 12). Figure 3b plots the de-trended series of percent monthly change in suicides after removal of detected autocorrelation parameters (MA 1, 12; I 12). This series exhibits no temporal pattern and serves as the exposure or independent variable of our test (Figure 3b).

**Figure 3:**
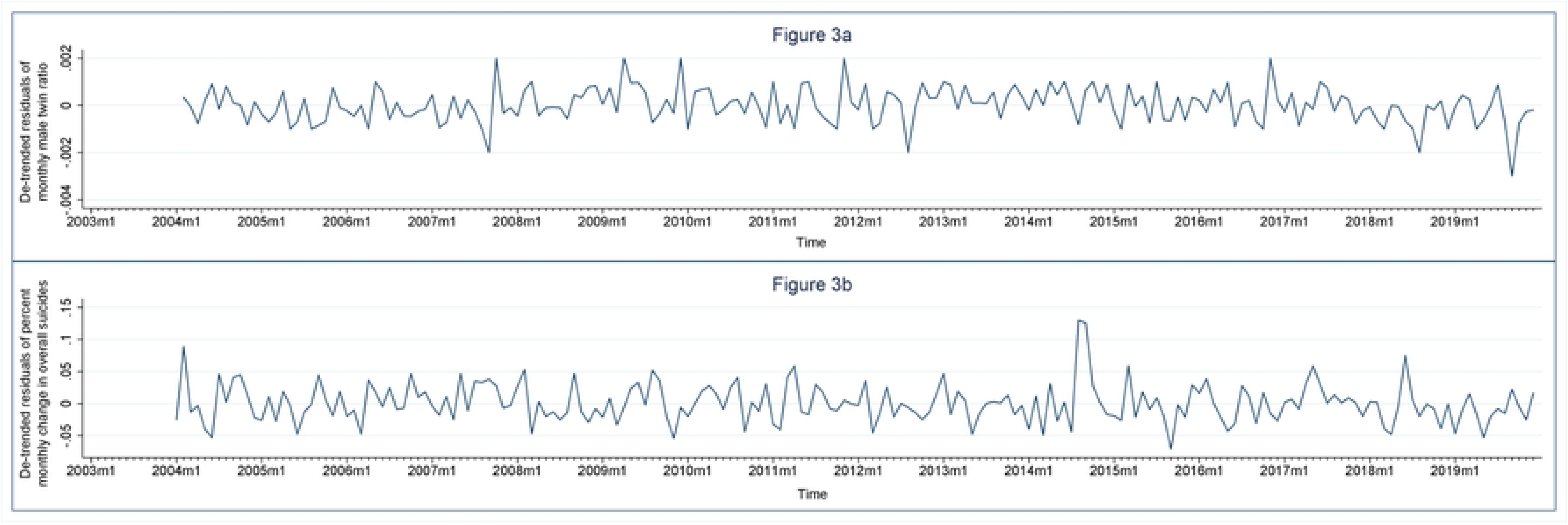
Residual series (after removal of autocorrelation) of {i) monthly male twin ratios (Figure 3a) and (ii) percent monthly change in overall suicides (Figure 3b), from January 2003 to December 2019, USA. Initial 12 observations lost to autocorrelation parameter modelling.

Table 2 shows the estimated coefficients for our time-series test model (corresponding to analytic step 3).

**Table 2:** Time-Series results for monthly male twin ratios from January 2003 to December 2019, as a function of exposure to de-trended residuals of percent monthly change in overall suicides and autocorrelation parameters.

| Variable | Coefficient | Standard Error |
| --- | --- | --- |
| Constant | 0.034 | 0.000*** |
| Autoregressive parameter: AR 1 | 0.210 | 0.076** |
| Autoregressive parameter: AR 12 | 0.666 | 0.058*** |
| Differencing | None |  |
| Exposure lag (de-trended residuals of percent monthly change in overall suicides) |  |  |
| 2 | 0.002 | 0.002 |
| 3 | 0.000 | 0.002 |
| 4 | 0.001 | 0.002 |
| 5 | 0.000 | 0.002 |
| 6 | -0.005 | 0.002** |

\*p < 0.05; two-sided test.
\*\*p < 0.01; two-sided test.
\*\*\*p < 0.001; two-sided test.

A 1 percent increase in overall suicides corresponds with a 0.005 unit decline in male twin ratio 6 months later (*p*= 0.004). Put another way, a one percent increase in overall suicides in a month (relative to the previous month) precedes a decline in 5 male twin live births per 1,000 male singleton live births 6 months later. This translates to a 15% decline in the average incidence of male twin births per 1000 male singleton live births (based on 33.4 average monthly male twin births per 1000 male singleton births in our sample; 5/33.4=15). Examination of residuals indicates absence of autocorrelation (Supplement Figure 2). Sensitivity tests support our original inference in that log-transformed series of monthly male twin ratio exhibits similar relation to de-trended residuals of percent monthly change in overall suicides at exposure lag of 6 months (coefficient = -0.137, *p* = 0.004) (Table 3).

**Table 3:** Time-Series results for log-transformed monthly male twin ratios from January 2003 to December 2019, as a function of exposure to de-trended residuals of percent monthly change in overall suicides and autocorrelation parameters.

| Variable | Coefficient | Standard Error |
| --- | --- | --- |
| Constant | -3.394 | 0.007*** |
| Autoregressive parameter: AR 1 | 0.211 | 0.076** |
| Autoregressive parameter: AR 12 | 0.670 | 0.057*** |
| Differencing | None |  |
| Exposure lag (de-trended residuals of percent monthly change in overall suicides) |  |  |
| 2 | 0.041 | 0.047 |
| 3 | 0.008 | 0.047 |
| 4 | 0.014 | 0.047 |
| 5 | 0.006 | 0.047 |
| 6 | -0.137 | 0.046** |

\*p < 0.05; two-sided test.
\*\*p < 0.01; two-sided test.
\*\*\*p < 0.001; two-sided test.

As a robustness check and to maintain comparability with prior work, we repeated our main test (from Table 2) with de-trended residuals of percent monthly change in female suicides (aged 15-49 years) as our exposure (Supplement Figure 3). Results from this robustness check support our main results in terms of exposure lag and statistically detectable decline in male twin ratio (Supplement Table 1). However, the relation between the outcome and exposure at lag 6 appears attenuated in magnitude (coefficient = - 0.0012, *p* = 0.014) suggesting a stronger relation between male twinning and changes in overall suicides, relative to male twinning and suicides among reproductive-aged females in the US (Supplement Table 1).

## Discussion

Volatility in collective optimism, or (conversely) collective despair, as indicated by national frequency of suicides, may precede *selection in utero* in a population ^13, 21^. Prior evidence from Sweden reports the presence of this relation with respect to extreme variations in female consumer confidence ^21^ and suicides among women aged 15-49 years ^13^. We extended this work to the US population and tested whether monthly changes in overall suicides predict changes in the ratio of male twin to male singleton live births—one sensitive indicator of *selection in utero*. Time-series test results indicate a 0.5% decline in male twinning per 1000 live-born male singletons, 6 months following an increase in overall suicides. We also observe a similar, albeit attenuated, relation between male twin live births and percent monthly change in suicides among 15-49 years old women. Our results align with prior work ^13, 21^ and suggest that the temporal patterning of suicides may correspond with male twin live births, which in aggregate may indicate a biological risk-averse response to volatility in collective optimism across varied populations.

Strengths of our study include the use of rigorous time-series modelling approaches that limit confounding from shared patterning (e.g. seasonality) across male twin births and suicides. In addition, we measure the residual series of percent monthly change in suicides in order to capture sudden perturbations (net of autocorrelation). This approach, we argue, provides a more valid test of *selection in utero* following acute changes in collective optimism, relative to observed monthly suicide counts. We also overcome a potential limitation of prior work by excluding female twin births from our outcome formulation ^13^. Rather, we examine the ratio of male twins to male singleton live births. In addition, our use of publicly available data from CDC WONDER allows independent replication and verification.

Moreover, we show that the theory of collective optimism may not only apply to suicides among reproductive-aged females ^13^, but to overall suicides in the US. The implications of this extension appear two-fold. First, our results suggest that male, in addition to female, suicides provide a sensitive gauge of collective optimism. Second, given that Catalano et al. (2020) used as a proxy for collective optimism suicides among females of child-bearing age – the only population in whom selection *in utero* operates – it remains possible that pregnant women (and, by extension, male twin live births) respond not to collective optimism but rather exposure to despair among one’s own sociodemographic group. The inverse relation we observe between male twin live births and suicide in males and females of all ages, in contrast, suggests that the mechanism by which humans signal the need for, and share optimism, appears more fundamental. Future work that tests the concordance of suicides to male twin births by age, sex, and racial/ethnic groups (e.g., the sensitivity of Black male twin births to suicides among Black women) can advance this hypothesis.

Limitations include that, as with most observational studies, we cannot eliminate the possibility of confounding from unobserved factors. Such a factor would (1) not exhibit any correlation with national trends in collective optimism, (2) exhibit simultaneous correlation with monthly changes in suicides and monthly male twin ratios at 6-month lag, and (3) exhibit orthogonality to seasonal patterning in suicides and male twin ratios. Whereas several phenomena such as economic recessions, political unrest, foreign or domestic terrorism and disease outbreaks may correspond with heightened *selection in utero* ^28–30^ and contemporaneously increase the incidence of suicide ^46^, we contend that such factors would also manifest as increased collective despair or diminished optimism.

Similar to prior work ^13^, another limitation of our study is that we do not distinguish between monozygotic versus dizygotic twins owing to non-reporting of this information in data provided by CDC WONDER. In addition, we cannot comment on whether collective optimism may correspond with increased incidence of vanishing twins (*in utero* absorption of a fetus by its twin, resulting in singleton live birth) ^47^, but we encourage future research to examine these relations when the appropriate population-level data become available. We also caution readers that the present study is strictly correlational in that we do not propose a causal link between increase in suicides and decline in male twinning. Our study, rather, suggests that ecological stressors that increase collective despair may extend beyond immediate effects on population-level psychiatric outcomes, into perinatal outcomes as well.

‘Deaths of despair’ comprise suicide, drug overdose, and alcohol use-related mortality ^1^. Although our study analyzed suicide, investigation into drug and alcohol related deaths may enhance the theoretical context of collective optimism^1^. Monthly incidence of psychiatric emergencies for mood and anxiety disorders and suicidal ideation/self-harm may also serve as measures of national mood. These indicators exhibit substantial variations across different regions in the US ^48, 49^, which may allow future research to conduct a detailed analysis of sub-national relations between regional optimism and male twin births in the US.

In addition to measures of psychiatric morbidity and mortality, indicators of risk-averse behavior may also serve as useful exposures for further tests of collective optimism ^21^. Analogous to Karasek et al (2015) who utilize a Swedish consumer confidence index to gauge risk aversion, surrogate measures such as new home ownership, home values and purchase of durable assets (e.g. automobiles) may reflect social optimism and willingness to invest in the future ^21^. Literature on one of the most widely studied ambient stressors-economic recessions-indicates that decline in consumption of sinful goods (alcohol, tobacco) and reduction in motor vehicular accidents may indicate increased risk aversion in a population ^50, 51^. This literature also reports decline in fertility as a potential risk-averse response to macroeconomic uncertainty ^52, 53^. We encourage future research to examine the relation between these proxy measures of collective optimism and *selection in utero* with respect to male twinning in the US.

Darwinian expectations from *selection in utero* would suggest that males born among cohorts with lower-than-expected male twin live births exhibit stronger survival characteristics ^24^. To explore potential changes in cohort fitness, re-tests of collective optimism and *selection in utero* may include examination of preterm births and early neonatal deaths among males ^54^, male-specific infant mortality ^55^, birth defects among live-born males ^56^, and incidence of other genetic conditions such as childhood cancers ^57^ among conception cohorts exposed *in utero* to greater-than-expected suicides ^22^. We expect these analyses to provide evidence of whether changes in collective optimism affect the survival characteristics of live-born cohorts, or if their relation to *selection in utero* against male twins diminishes beyond parturition.

## Data Availability

All data files on national monthly suicides and male twin births are publicly available from the CDC WONDER database

https://wonder.cdc.gov/

---

Supplement Figure 1: Plot of percent monthly change in suicides among females aged 15-49 years in the US, from January 2003 to December 2019.

Supplement Figure 2: Plot of autocorrelation function (ACF) up to 12 month lags of monthly male twin ratio after specifying autoregressive parameters at lags 1 and 12 (AR 1, 12).

Supplement Figure 3: Residual series (after removal of autocorrelation) of percent monthly change in suicides among women aged 15-49 years, from January 2003 to December 2019, in the US. Initial 12 observations consumed in autocorrelation parameter modelling.

**Supplement Table 1:** Time-Series results for monthly male twin ratios from January 2003 to December 2019, as a function of exposure to de-trended residuals of percent monthly change in suicides among females aged 15-49 years and autocorrelation parameters.

| Variable | Coefficient | Standard Error |
| --- | --- | --- |
| Constant | 0.034 | 0.000*** |
| Autoregressive parameter: AR 1 | 0.076 | 0.077 |
| Autoregressive parameter: AR 12 | 0.688 | 0.052*** |
| Differencing | None |  |
| Exposure lag (de-trended residuals of percent monthly change in suicides among females aged 15-49 years) |  |  |
| 2 | 0.0000 | 0.0005 |
| 3 | 0.0000 | 0.0006 |
| 4 | 0.0002 | 0.0007 |
| 5 | -0.0001 | 0.0006 |
| 6 | -0.0012 | 0.0005* |

\*p < 0.05; two-sided test.
\*\*p < 0.01; two-sided test.
\*\*\*p < 0.001; two-sided test.

